# Harnessing the Open Access Version of ChatGPT for Enhanced Clinical Opinions

**DOI:** 10.1101/2023.08.23.23294478

**Authors:** Zachary M Tenner, Michael Cottone, Martin Chavez

**Affiliations:** New York University Grossman School of Medicine, Mineola, New York, USA; Division of Maternal-Fetal Medicine, Department of Obstetrics Gynecology, New York University Langone Hospital-Long Island, New York University Grossman Long Island School of Medicine, Mineola, New York, USA

**Author notes:** **<u>Corresponding Author:</u>** Zachary Tenner – (ZT).

## Abstract

With the advent of Large Language Models (LLMs) like ChatGPT, the integration of AI into clinical medicine is becoming increasingly feasible. This study aimed to evaluate the ability of the freely available ChatGPT-3.5 to generate complex differential diagnoses, comparing its output to case records of the Massachusetts General Hospital published in the New England Journal of Medicine (NEJM). Forty case records were presented to ChatGPT-3.5, with prompts to provide a differential diagnosis and then narrow it down to the most likely diagnosis. Results indicated that the final diagnosis was included in ChatGPT-3.5’s original differential list in 42.5% of the cases. After narrowing, ChatGPT correctly determined the final diagnosis in 27.5% of the cases, demonstrating a decrease in accuracy compared to previous studies using common chief complaints. These findings emphasize the need for further investigation into the capabilities and limitations of LLMs in clinical scenarios, while highlighting the potential role of AI as an augmented clinical opinion. With anticipated growth and enhancements to AI tools like ChatGPT, physicians and other healthcare workers will likely find increasing support in generating differential diagnoses. However, continued exploration and regulation are essential to ensure the safe and effective integration of AI into healthcare practice. Future studies may seek to compare newer versions of ChatGPT or investigate patient outcomes with physician integration of this AI technology. By understanding and expanding AI’s capabilities, particularly in differential diagnosis, the medical field may foster innovation and provide additional resources, especially in underserved areas.

## Introduction

Research and speculation regarding the integration of artificial intelligence (AI) into physician reasoning has been ongoing since the 20^th^ century. In 1987, Schwartz et al., asserted that “major intellectual and technical problems must be solved before we can produce truly reliable [healthcare] consulting programs”(1). Models of clinical problem solving have been described for years, but it is only recently that technology has advanced sufficiently to investigate the role of AI in clinical medicine. OpenAI’s ChatGPT (Generative Pre-trained Transformer), one of the world’s first widely used Large Language Models (LLM), uses billions of parameters to generate user-informed text. In the healthcare sector, this generative artificial intelligence encompasses a range of medical knowledge that can be tailored to the user’s needs, from assisting medical students with United States Medical Licensing Exam (USMLE) questions to creating next-generation sequencing reports with treatments options for attending oncologists (2, 3). Upon its release, professionals began assessing the value of ChatGPT by pushing its limits within medical knowledge; however, it is imperative to explore ChatGPT’s role in patient care to best demonstrate and provide direction for how health professionals will work with AI as technology develops (4, 5).

ChatGPT has distinguished itself by achieving passing scores on the USMLE examination, equivalent to those of a third-year medical student (2). This accomplishment opens the gates for potential applications of the model for interactivity in medical school and an overall tool to support clinical thinking. Radiology and pathology have received significant attention in AI research, through efforts of enhancing LLMs to better understand images and detect cancers. Despite no specific training within either subject, “ChatGPT nearly passed a radiology board-style examination without images,” and demonstrated accuracy in “[solving] higher-order reasoning questions in pathology” (6, 7). Ali et. al. identified ChatGPT’s ability to perform at high rates on the neurosurgery oral boards examination preparation while emphasizing the limitation in using multiple-choice examinations to assess a neurosurgeon’s expertise in patient management (8). Although ChatGPT has proven effective in choosing from a list of options, the role of LLMs in clinical management has been highlighted as area requiring further research.

Mirroring the progression of a medical student, the next logical step is to evaluate the chatbot’s ability to come up with differential diagnosis. These are fundamental to clinical medicine, and the proficiency of ChatGPT in generating medically rational differential diagnoses remains largely unexplored. Hirosawa et al. determined that ChatGPT can successfully create comprehensive diagnosis lists for common chief complaints (9). Additionally, Rao et al. assessed ChatGPT’s ability to generate differential diagnosis for routinely encountered in healthcare settings and found, “the LLM demonstrated the highest performance in making a final diagnosis with an accuracy of 76.9%” (10). Previous research has done a great job of assessing ChatGPT’s ability to pass multiple-choice exams and provide differential diagnosis for standard chief complains with high accuracy; however, the generalizability of ChatGPT to more complex clinical scenarios must be examined (11).

To truly assess the potential of AI and LLMs in complex medical reasoning, we tested the ability of the freely available ChatGPT-3.5 to provide differentials on case records of the Massachusetts General Hospital published in the New England Journal of Medicine (NEJM). Our research further evaluates the chatbot languages by using clinical case reports that have been identified by the journal to establish novel medical or biological understanding. Launched in 2022, ChatGPT-3.5 has a knowledge cutoff date of September 2021; therefore, we were able to examine ChatGPT’s ability to use clinical reasoning to diagnose 2022 case reports, rather than rely on its search function to locate published articles. The aim of this study is to evaluate the freely available ChatGPT-3.5’s proficiency in generating complex differential diagnoses. We intend to compare the chatbot’s complete diagnosis list and final diagnosis against the published differential diagnosis for the NEJM case reports. We hypothesize that the percentage of differential diagnoses generated by ChatGPT-3.5 will match the NEJM final diagnosis for the case reports about 50% of the time. By elucidating ChatGPT’s potential in offering differential diagnoses, we propose future clinical problem-solving cases to consider utilizing AI as an augmented clinical opinion.

## Methods

Forty case records from the Massachusetts General Hospital published in the New England Journal of Medicine (NEJM) in 2022 were presented to ChatGPT-3.5. All text prior to the Differential Diagnosis headline was included (excluding figures). ChatGPT was first prompted to, “Provide a differential diagnosis from the following clinical case.” After generating a complete list of differential diagnoses, we asked ChatGPT, “Can you narrow down the differential to the most likely diagnosis?” From these prompts, we recorded whether the final diagnosis, as referenced in the NEJM, was included in the complete differential diagnosis list and whether ChatGPT’s “most likely diagnosis” aligned with the final diagnosis noted in NEJM.

## Results

Of the 40 cases presented to ChatGPT-3.5, 23 cases (57.5%) were not considered in its’ original differential list. The average length of the original differential list produced by ChatGPT was 7.4±2.2 possible diagnoses with a high of 12 and a low of 3. The length of the differential appeared random. In 17 cases (42.5%) ChatGPT did include the final diagnosis in its’ original differential list. After narrowing down its’ differential list, ChatGPT correctly determined the final diagnosis in 11 cases (27.5%) and eliminated the correct diagnosis in 6 cases (15%). These results are presented in Figure 1.

**Figure 1.**
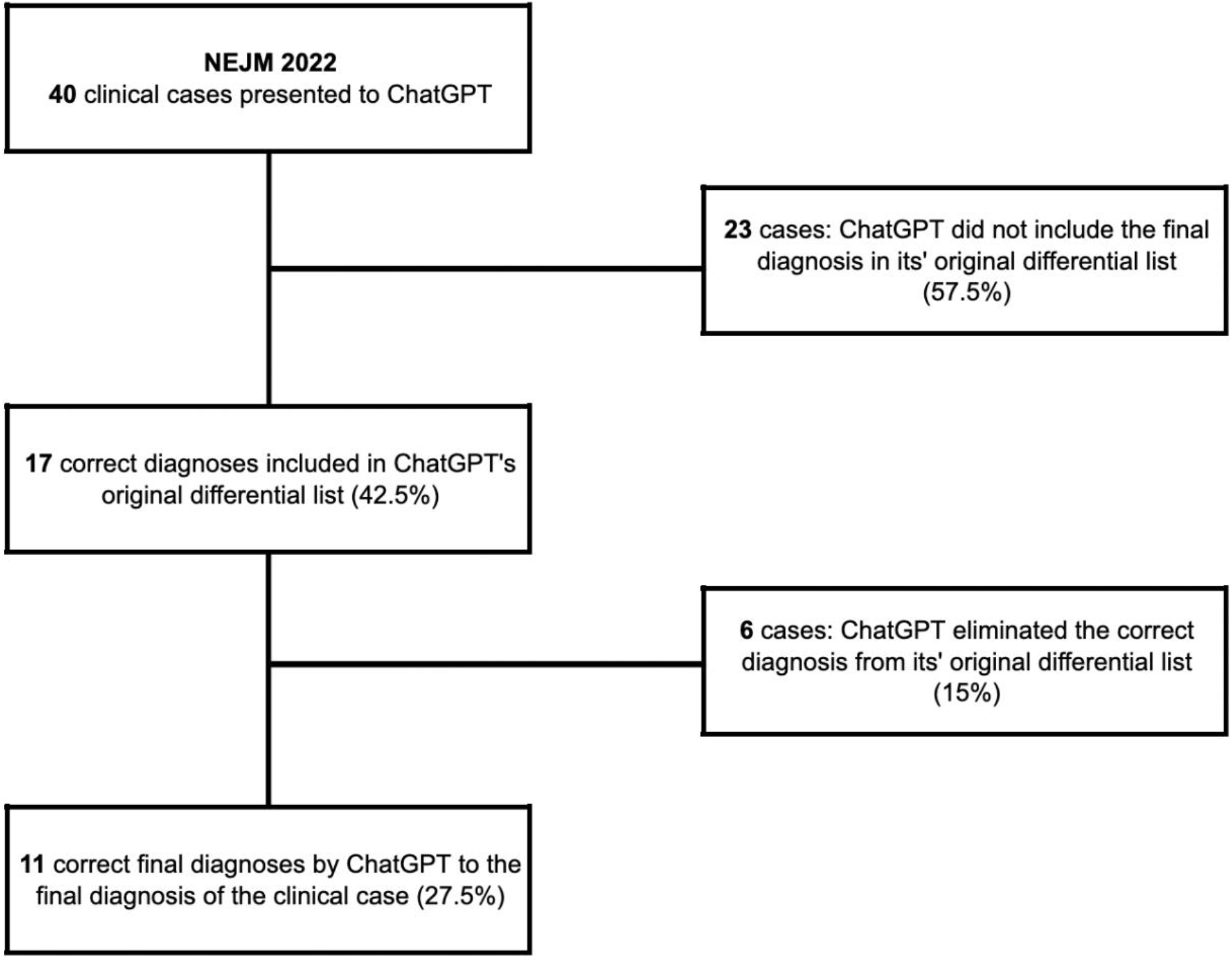
Flowchart of the 40 case records of the Massachusetts General Hospital that were published in the NEJM after being presented to ChatGPT.

## Discussion

The role of generative AI and LLMs in clinical medicine is a rapidly growing area of research. Assessing the potential and limitations of ChatGPT (v3.5) within the scope of patient care is essential to determine how and where it can best be utilized. We decided to focus on the complimentary version of ChatGPT since we wanted the largest possible audience to have access to this technology. We presented 40 case records of the NEJM to ChatGPT to further research LLMs role in healthcare and study its success rates in producing differential diagnoses of complex patient presentations. ChatGPT reached the correct differential diagnosis 27.5% (11/40) of the time. The differential list accuracy of ChatGPT when presented with clinical vignettes of common chief complaints has been reported to be over 80% (9).That accuracy dropped by over 50% when we increased the number of clinical cases using NEJM case reports. Establishing baseline limitations of ChatGPT allows for future comparisons of its growth and development and ensures cautious use in patient care. Furthermore, it can provide insight to how to best adjust ChatGPT’s setting to better identify the categories for which it receives the highest score.

OpenAI has begun to introduce plugins that provide real-time access to data that enhance the program’s capabilities. Physicians and other healthcare workers will soon be practicing in a world where the latest research journals and electronic medical records are directly linked to these Chat-like software. With these new additions coming to ChatGPT, we expect ChatGPT to continue growing in its ability to develop differential diagnoses. As a result, it is ever so important to the field of medicine for this information to be better understood. In both primary care and specialty settings, AI offers a new medium for physicians to foster new ideas, consider new diagnoses, and consult with a “colleague” when one may not be available, such as in rural settings (12).

Future studies may look to expand from our baseline findings. How do newer versions of ChatGPT compare to ChatGPT-3.5? Do patients have better outcomes when their physician implements ChatGPT into their care? These questions, and many more, are to be elucidated with further experimentation; however, before ChatGPT does become a new tool within a physician’s practice, we must first continue to define and describe abilities to ensure AIs safe use and appropriate reliance. We strongly advocate for technology companies to consistently offer complimentary versions of generative artificial intelligence. Doing so not only maximizes its utilization but also fosters innovation, particularly in the field of medicine.

## Data Availability

All relevant data are within the manuscript.

